# Pretrained Language Models for Semantics-Aware Data Harmonisation of Observational Clinical Studies in the Era of Big Data

**DOI:** 10.1101/2024.07.12.24310136

**Authors:** Jakub Dylag, Zlatko Zlatev, Michael Boniface

**Affiliations:** IT Innovation Centre, School of Electronics and Computer Science, University of Southampton, Southampton, United Kingdom

## Abstract

**Background:** In clinical research, there is a strong drive to leverage big data from population cohort studies and routine electronic healthcare records to design new interventions, improve health outcomes and increase efficiency of healthcare delivery. Yet, realising these potential demands requires substantial efforts in harmonising source datasets and curating study data, which currently relies on costly and time-consuming manual and labour-intensive methods.

**Objectives:** We evaluate the applicability of AI methods for natural language processing (NLP) and unsupervised machine learning (ML) to the challenges of big data semantic harmonisation and curation. Our aim is to establish an efficient and robust technological foundation for the development of automated tools supporting data curation of large clinical datasets.

**Methods:** We assess NLP and unsupervised ML algorithms and propose two pipelines for automated semantic harmonisation: a pipeline for semantics-aware search for domain relevant variables and a pipeline for clustering of semantically similar variables. We evaluate pipeline performance using 94,037 textual variable descriptions from the English Longitudinal Study of Ageing (ELSA) database.

**Results:** We observe high accuracy of our Semantic Search pipeline with an AUC of 0.899 (SD=0.056). Our Semantic Clustering pipeline achieves a V-measure of 0.237 (SD=0.157), which is on par with leading implementations in other relevant domains. Automation can significantly accelerate the process of dataset harmonization. Manual labelling was performed at a speed of 2.1 descriptions per minute, with our automated labelling increasing speed to 245 descriptions per minute.

**Conclusions:** Our study findings underscore the potential of AI technologies, such as NLP and unsupervised ML, in automating the harmonisation and curation of big data for clinical research. By establishing a robust technological foundation, we pave the way for the development of automated tools that streamline the process, enabling health data scientists to leverage big data more efficiently and effectively in their studies, accelerating insights from data for clinical benefit.

## Introduction

Clinical research plays a vital role in advancing medical knowledge and improving patient care. Traditionally, clinical studies have employed randomized controlled experiments and prospective studies. These approaches can be time-consuming, resource-intensive, and may not always be feasible in certain clinical research contexts. In recent years, observational retrospective clinical studies have emerged as valuable alternatives that offer notable advantages in terms of cost and efficiency and can still yield valid results [1].

One significant catalyst behind the rise of observational retrospective clinical studies is the availability of extensive cohort and routine clinical practice databases, including notable examples like the English Longitudinal Study of Ageing (ELSA), Clinical Practice Research Datalink (CPRD), and Secure Anonymized Information Linkage (SAIL). These databases are characterized by their large size and heterogeneity, collectively forming what is commonly referred to as *big data* in the field of data science.

Leveraging healthcare big data offers great potential for discovering insights into diverse clinical questions exploring the complexities of multiple long-term conditions (MLTCs), designing new interventions to improve healthcare outcomes and improving the quality and efficiency of healthcare delivery [2]. However, exploiting this potential requires significant effort in harmonising source datasets and curating the study data [3]. In observational studies, such as the Cluster-AIM study for development and validation of population clusters for integrating health and social care for patients with MLTCs [4], datasets must be curated from various cohort study databases or routine healthcare data databases. Such studies have complex and multi-faceted domains with 10s of thousands of variables to be curated for the specific research task at hand. The process of datasets harmonisation and study data curation encompasses several crucial steps. This includes defining the domains and sub-domains of interest, identifying relevant variables within these sub-domains, identifying the equivalent variables, and extracting the necessary data from the databases. Working with big data poses sizeable challenge particularly during the variable identification process as the datasets can feature extensive numbers of domains and sub-domains, increasing the difficulty of variable selection and study dataset harmonisation within available time constraints [5,6].

Furthermore, the absence of standards, frameworks, and journal requirements for the reporting and sharing of data harmonisation outcomes results in loss of resources, time, and effort [7]. Often, variable names and descriptions are ambiguous and inconsistent across datasets which increases the difficulty of dataset harmonisation [8].

Given the vastness of information in big data, comprising of thousands of variables and recorded over extensive periods of time, researchers face the daunting task of sifting through large collections of variables’ descriptions to identify the ones pertinent to their study objectives. This process demands considerable time and effort, often extending over many weeks and months. Researchers must meticulously draft sub-domain descriptions, identify relevant search terms, conduct thorough searches within the exceptionally large collections of variable descriptions, and review and select the variables that align with the defined sub-domains of interest.

In the current manuscript, our work focuses on the research and validation of machine learning (ML) technologies to facilitate the creation of automated tools that aid in the harmonisation of datasets and the curation of research data for observational studies from healthcare big data sources. We explore advancements in the fields of Natural Language Processing (NLP) and unsupervised ML techniques. By utilising these technologies, we demonstrate how the variable identification process can be streamlined, reducing the time and effort required for dataset curation. To evaluate the efficacy of the selected ML methods, we employ the ELSA datasets, specifically targeting the study of social care needs for people living with MLTCs. The domain was selected due to the complexity of variables and relevance to the Cluster-AIM study.

The rest of the manuscript is organized as follows: The methods section provides a description of the data utilized in the current study, the proposed data harmonisation and curation pipelines, the technologies employed in the pipelines, and the methods used to evaluate their performance. In the results section we present the corresponding evaluation results, which are then further analysed. The manuscript concludes with a discussion on the implications and potential applications of our findings, highlighting the benefits of automated tools for dataset harmonisation and curation in observational studies utilizing healthcare big data.

## Methods

In his work, Bosch-Capblanch [8] defines three key characteristics necessary for the harmonization of variables: a unique identifier, a semantically identical description, and consistent statistical metrics for its values. Cunningham et al. [9] further define semantic harmonization as the process of collating this data into a singular consistent logical view. Although harmonisation and curation tools, such as BiobankConnect software [10], SORTA [11] and DataSHaPER [12] exist, their operation is underpinned by expert crafted ontology- and schema-based data annotation, which are difficult to create. Simpler rule-based approaches have also been employed but these rely on variable names similarity and are not general [8]. An alternative that can overcome these challenges is the use of data-driven Artificial Intelligence (AI) and Machine Learning (ML) algorithmic approaches [9]. Using techniques such as Natural Language Processing (NLP) and unsupervised learning we demonstrate tools supporting semantic data harmonisation and curation. We evaluate the performance in terms of accuracy and time savings of two semantic harmonisation automation pipelines: (1) Semantic Search for domain-relevant variables and (2) Semantic Clustering semantically similar variables.

### Evaluation Dataset

We use the English Longitudinal Study of Ageing (ELSA) [13] datasets to evaluate the semantic data harmonisation process. The ELSA study surveyed households with at least one adult aged over 50 with the aim to gain insight into all aspects of the UK’s ageing population. The Study was conducted as a series of 10 stages, commencing in 1998 with the most recent stage finishing in 2019. Each Wave took place 2 years after the previous, with the same participants surveyed, subject to consent and other extenuating circumstances. A total of over 18,000 people participated in the study, with consistent population of over 8,000 throughout the later 9 waves. The sample is based on respondents in the Health Survey for England (HSE), which annually surveys health and lifestyle changes. A variety of data collection methodologies were used including, Face-to-Face Interviews, Assisted Measurements (both Clinical and Physical) and Questionnaires (both paper-based and web-based). Local area data can enable data linkage with consensus data concerning income, education, and employment.

Although attempts have been made by Lee et al [14] to harmonise the ELSA datasets, not all available data has been incorporated. Additionally, no use of harmonisation tools is reported. In ELSA, 94,037 variables are recorded across 67 tabular files, leading to significant difficulties when navigating and analysing the datasets. This complexity makes ELSA an ideal use case for testing the proposed semantic harmonisation methodology.

The number of variables across all waves in the ELSA study can be seen in Figure 1. A significant portion of variables across the ELSA datasets for waves 1-9 capture longitudinally the same information but don’t have consistent naming between waves. Following the Bosch-Capblanch definition for harmonization of variables [8] we perform ELSA identifier-level harmonization by matching variable identifiers in a case-insensitive manner. This initial step ensures that variables with the same identifiers are recognized and treated as identical, despite potential variations in case. The identifier harmonisation eliminated variable identifier duplication, observing a reduction from 94,037 variables to 22,402 unique variables.

**Figure 1:**
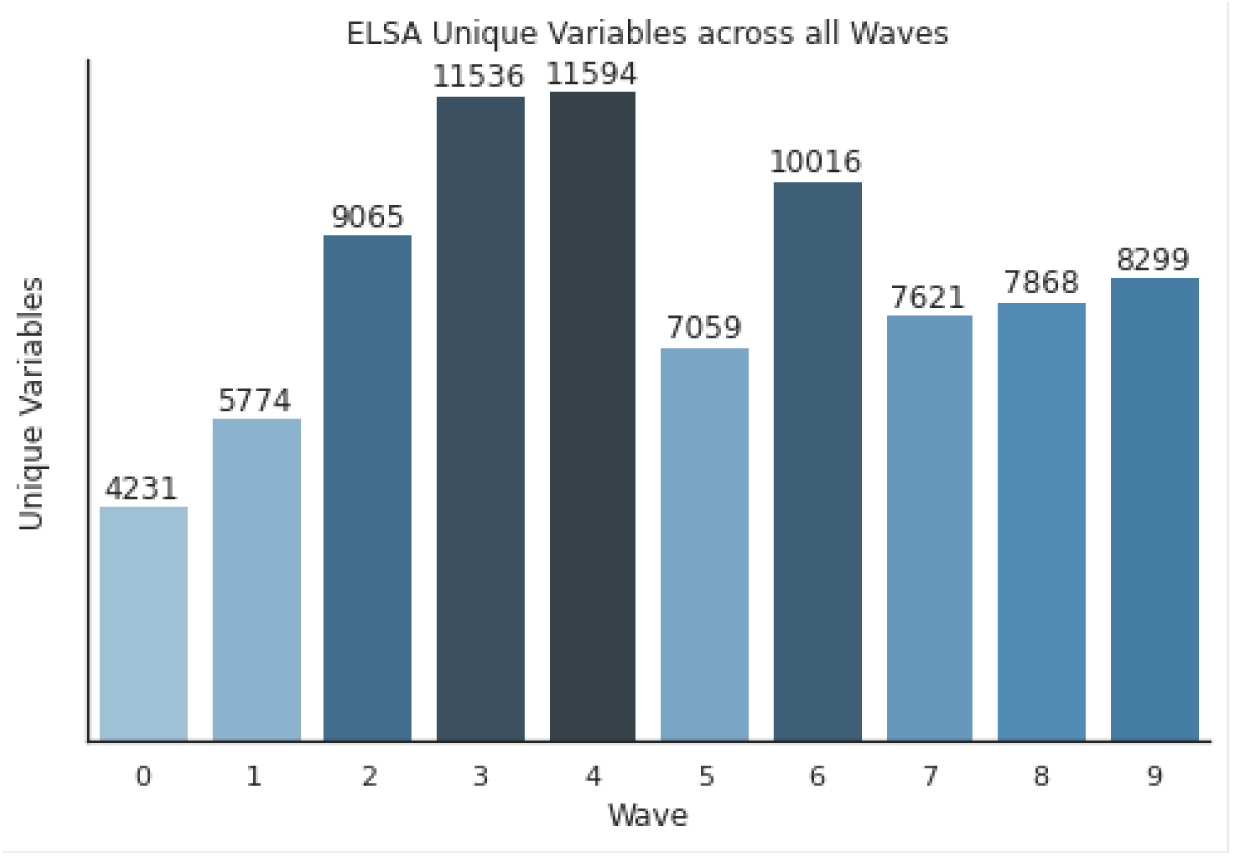
Number of ELSA variables in each wave.

### Semantic Harmonisation Methodology

The focus of our study is on the semantic analysis of variable descriptions to identify semantically identical variables using NLP and ML technologies. We discuss the state-of-the-art semantics-aware text embedding technologies that underpin our approach. We then detail the design and implementation of the two semantic harmonization pipelines: 1) Semantic Search to identify domain-relevant variables and 2) Semantic Clustering of similar variables.

#### Efficient Semantics-Aware Text Embedding

We investigate NLP technologies that can efficiently generate text embeddings that capture semantic context for our harmonisation pipelines. NLP embeddings (i.e., dense vector representations) have gained prominent use in medical research for analysing unstructured textual data from Electronic Healthcare Records (EHR), Intensive Care Units (ICU), social media and Scientific Literature [15,16]. Embedding models are trained in an unsupervised manner, capturing knowledge from large unlabelled corpuses in high dimensional vector spaces. These embeddings can be leveraged in semantics-aware clustering and search tasks.

Numerous methods of Sentence Embeddings have previously been proposed. Skip-Thought [17] trains an encoder-decoder gated recurrent unit (GRU) architecture to predict surrounding sentences from a given passage using an unsupervised methodology. By utilising the encoder, a latent space of semantically similar sentences is created, enabling use in semantic similarity tasks. Universal Sentence Encoders (USE) [18] improve upon Skip-Thought by introducing a transformer network for significant performance gains at the expense of model complexity, computation time and memory usage. Contextual embeddings aware of ordering and identity of each word are first computed, and subsequently summed at each word position into a fixed size 512-dimensional vector. The encodings are designed to be general-purpose and applicable to a wide range of domains. Chen et al [16] utilises USEs within the healthcare domain to find similar sentences in EHR. However, testing on the BEIR dataset [19] indicates subpar performance on when compared to other Neural based methods.

Bidirectional Encoder Representations from Transformers (BERT) models [20] are pre-trained transformer network producing contextual embeddings. Words are tokenized using WordPiece [21] with a 30,000 token vocabulary, after which 12 layers of multi-head attention are applied and passed to a simple regression function. RoBERTa demonstrated further improvements by adapting the training process by tuning hyperparameters and expanding training set sizes. Although BERT based models can be adapted to embed sentences by iterative processing of singular words, it is limited to a pre-determined fixed sized sentence length, restricting comparison performance and increases storage requirements. Sequences of BERT word embeddings may be averaged, into a singular sentence vector [22,23] however this results in significant performance degradation.

Sentence-BERT (SBERT) [24] models have demonstrated good performance in Semantic Textual Similarity (STS) tasks, with semantically meaningful embeddings. It can map textual sentence input, up to 250 words in length, to a single fixed size vector. A modification is made of BERT architecture using Siamese, Triplet networks and subsequent pooling [20]. A cosine similarity objective function [24] is utilised to calculate similarity between processed sentences. Other metrics, such as the dot product, have been shown to outperform cosine similarity on specific datasets, however on average cosine similarity has marginally better performance [19].

We leverage the SBERT architecture to underpin our semantic data harmonisation and curation solutions. We analyse and compare four pretrained SBERT-based language models to empirically investigate the impacts of model size and training set domain on harmonisation performance. These four models are MiniLM, MPNet, Sentence-T5-xxl and BioLinkBERT, and their specific training details are described below.

#### MiniLM

MiniLM [25] is proposed by Wang et al and implements a SBERT architecture [24]. The model compresses large Transformer models into smaller, more efficient models through deep self-attention distillation. Leveraging subsequent development by Reimers et al [26] the MiniLM model was adapted to just six layers with an embedding vector size of 384. This results in the fastest inference times of 14200/sec on a V100 graphics processing unit (GPU). Training used 100 thousand steps on a tensor processing unit (TPU) v3.8, upon 1.17 billion sentence pairs, with majority from Reddit Comments [27], S20RC [28], WikiAnswers [29] and PAQ [30].

#### MPNet

MPNet [31] by Song et al improves upon the BERT [20] and SBERT pretraining methods by reducing positional discrepancies and leveraging dependencies amongst all tokens in a sentence through permutated language modelling. Further fine-tunning of MPNet has resulted in the creation of all-mpnet-base-v1 [32], pretrained on 1.1 billion sentence pairs as with MiniLM. This model has increased complexity, with 768-dimensional embedding space, slowing inference to 2800/sec on V100 GPU.

#### Sentence-T5-xxl

Text-to-Test Transfer Transformer (T5) introduced by Raffel et al [33] excels in a variety of NLP tasks by leveraging Colossal Clean Crawled Corpus [34] and harnessing transfer learning. Ni et al. [35] scaled up the T5 model to 11 billion parameters and incorporated an SBERT architecture to develop the Sentence-T5-xxl model. Sentence-T5-xxl retains state-of-art performance in sentence embedding tasks, with 768 dimensional embeddings, however at the expense of very slow inference (50/sec on V100 GPU). The model is trained on a corpus of two billion question-answer pairs from various online communities as well as the Stanford Natural Language Inference (SNLI) dataset [36].

#### BioLinkBERT

Yasunaga et al propose the LinkBERT [37] pretraining method which leverages links between documents, viewing a text corpus as a graph of documents and in so creating document contexts. This approach is especially relevant for the pretraining of domain specific models. BioLinkBERT is a pretrained language model using LinkBERT on PubMed to achieve state of the art performance in BioNLP tasks such as BioASQ [38] and USMLE [39]. The model uses a 512-dimensional embedding space and has comparable inference times to MPNet.

#### Language models vector space comparison

To get an insight on the models’ vector spaces, we computed and plotted the cosine distance distributions of the embeddings for all variable descriptions in our datasets – see Figure 2. The plot indicates important similarities and differences in the vector spaces of the four models. MiniLM (M = 0.869, SD = 0.142) and MPNet (M = 0.856, SD = 0.133) have similar distributions. T5 (M = 0.346, SD = 0.055) and BioLinkBERT (M = 0.189, SD = 0.067) have a significantly lower mean and denser distribution. The wider cosine distance distribution of MiniLM and MPNet compared to T5 and BioLinkBERT provides for higher discrimination ability in the downstream tasks.

**Figure 2:**
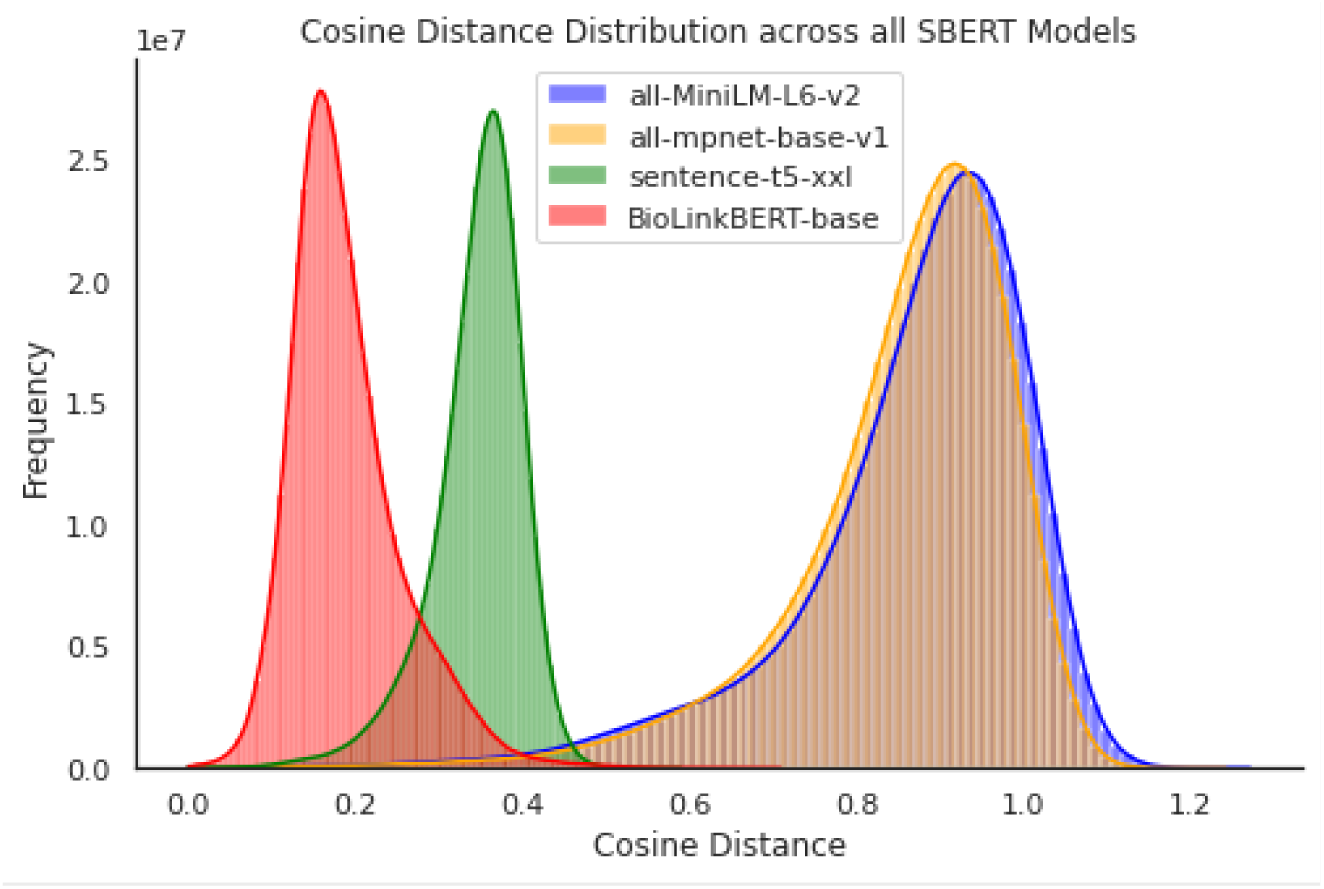
Illustration of distributions of cosine distances between normalized embedding vectors of each variable description across all SBERT models.

### Semantic Search for Domain-relevant Variables

Semantic harmonization is the process of collating data into a singular consistent logical view [9]. Often this logical view is the collation of variables relevant to domains of interest. Semantic Search can automate suggestion of variables within a domain.

Guha et al [40] introduces Semantic Search methodologies for improved web search results on the semantic web. Unlike previous approaches that merged textual and semantic information into single search indexes, this study uses inverted indexes for searching by textual content, contrasting with forward indexes, which fetch information using unique identifiers.

Traditional keyword-based retrieval models require explicit observation of search terms, therefore increasing index size and total query time. In contrast, neural embedding-based methods alleviate these inefficiencies by utilising a unified, both textual and semantic, embedding space [41]. For an instance, word embeddings have seen success in extending full text search for legal document collections [42].

In the current work we propose a neural embedding-based solution to automating semantics-aware search for variables relevant to given domain of interest. The solution enables the user to specify a phrase, whose embedding will be compared against all variable description embeddings, enabling the closest matches to be selected. This significantly reduces the time taken for variable selection, as well as improving performance over basic approaches such as keyword search, by leveraging semantic contexts. Based on our analysis of efficient semantics-aware text embedding technologies, we utilise the SBERT model architecture and evaluate the MiniLM, MPNet, BioLinkBERT and T5-XXL pretrained model.

As illustrated in Figure 3, we incorporate the SBERT model into the proposed Semantic Search pipeline. Embeddings of variable meta-data descriptions are precomputed, enabling the use of efficient Semantic Search methods. We use the cosine similarity function to compare qualitative domain specificphrase embeddings to all variable embeddings. Although other metrics, such as the dot product, are also appropriate, it has been shown that cosine distance has the best performance on average [19].

**Figure 3:**
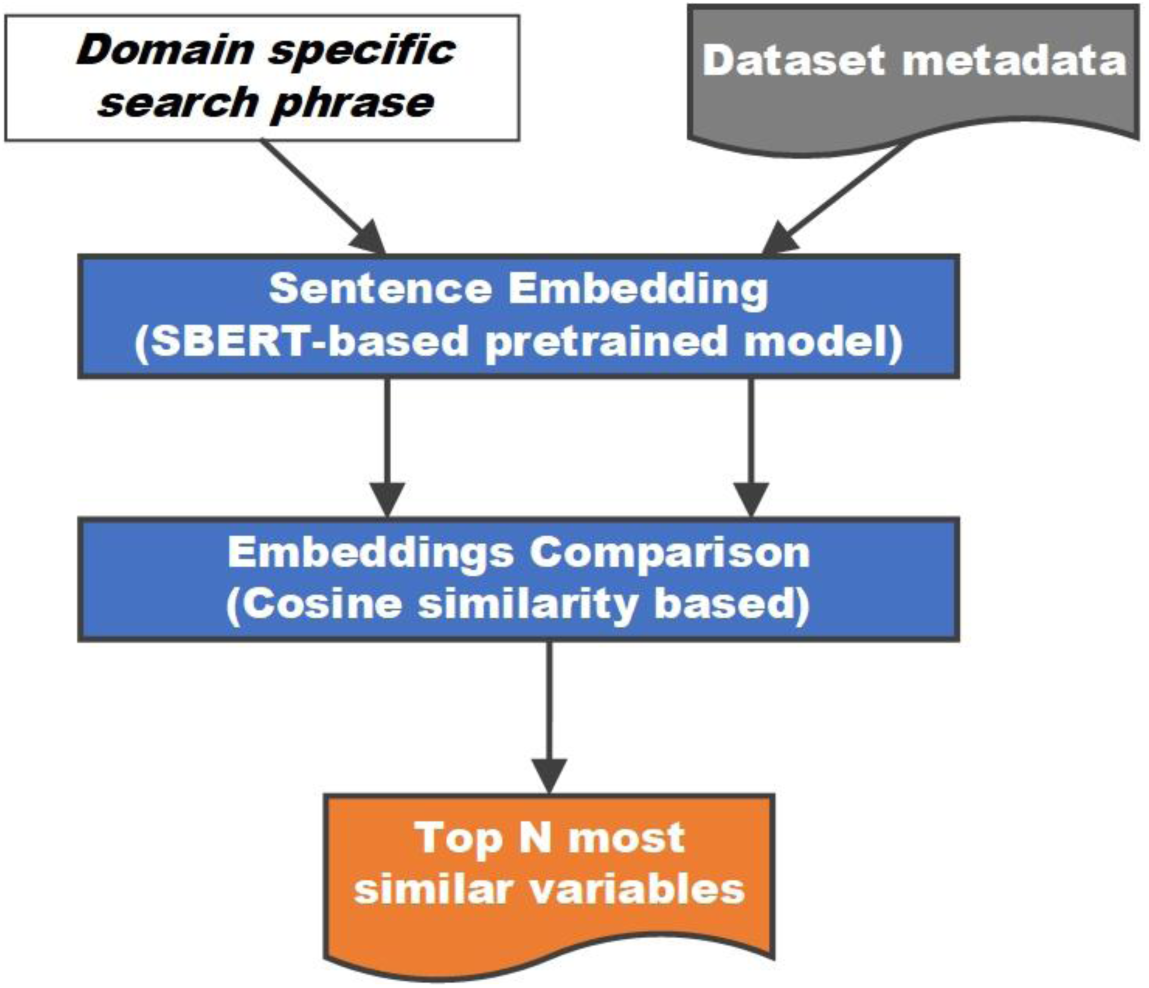
Implemented pipeline processes for Semantic Search of variable descriptions.

Finally, to select the domain-relevant variables, the proposed pipeline outputs the top N descriptions with the largest similarity to the search phrase can be chosen. An alternative to this current functionality could be to apply a thresholding function on the distance of the variables’ embeddings from the search phrase embedding. However, as presented in Figure 2, various models have varying sparsity of embeddings and therefore thresholds need to be appropriately adapted for each model.

### Semantic Clustering of Variables into Domains

Building on the pipeline for identifying variables relevant to a specific domain of interest, we propose a new pipeline for the unsupervised grouping of variables into semantically cohesive domains. We base this pipeline on unsupervised ML methods for dimension reduction and clustering to enable a fully automated grouping of semantically similar variables based on the sentence embeddings of the variable descriptions in the dataset metadata. Figure 4 depicts the pipeline for unsupervised variable domain clustering which, in addition to the text embedding algorithm, incorporates an algorithm for dimensionality reduction of the high dimensional embedding space and an algorithm for clustering. Variables within the same cluster are semantically similar and are harmonised together in the same domain.

**Figure 4:**
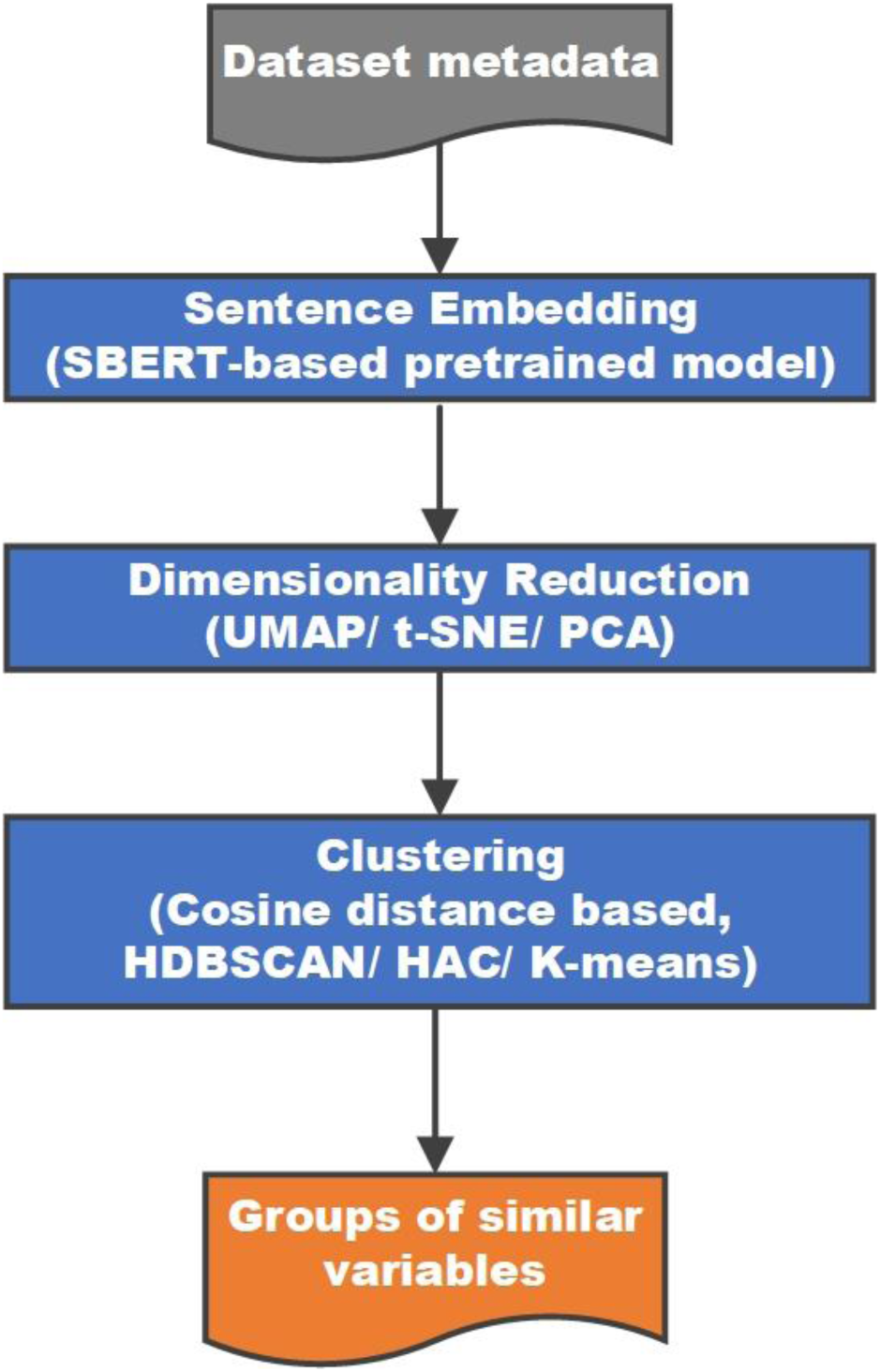
Implemented pipelines processes for unsupervised clustering of variable descriptions.

Previous efforts have been made to cluster embeddings of supervised models, with varying levels of success. Nikifarjam et al. [43] embedded short form tweets using Word2Vec [44], clustered these using K-means, after which a Conditional Random Fields classification model was trained upon. Xu et al. [45] uses K-means to cluster denseneural embeddings with a unique Convolutional Neural Network model. Bodrunova et al. [46] uses Hierarchal Agglomerative Clustering to group Universal Sentence Encoder embeddings, with the addition of Markov Stopping moment to choose optimal number of clusters. Similarly, An et al. [47] uses a range of both static and dynamic Sentence Embeddings, which are clustered with k-means at into a specified number of groups by Spatial Histogram analysis. Gupta et al. [48] makes the insight that lowering embedding dimensionality previous to clustering using an Encoder-Decoder Model, improves clustering performance.

The above unsupervised clustering algorithms require a pairwise dis-similarity to be computed for every combination of description embeddings. As stated previously, we use cosine similarity for the comparison of the SBERT embeddings. Cosine similarity is converted to cosine distance by the following simple conversion 𝑐𝑜𝑠𝑖𝑛𝑒 𝑑𝑖𝑠𝑡𝑎𝑛𝑐𝑒 = 1 − 𝑐𝑜𝑠𝑖𝑛𝑒 𝑠𝑖𝑚𝑖𝑙𝑎𝑟𝑖𝑡𝑦, as the clustering algorithms use a distance measure. Furthermore, embedding vectors are normalised prior to cosine distance calculations to ensure consistency between various embedding models.

For the pipeline in Figure 4 we compared three dimensionality reduction algorithms, namely PCA, t-SNE and UMAP, and also three clustering algorithms, namely K-means, Hierarchical Agglomerative Clustering and HDBSCAN.

#### Dimensionality Reduction Algorithms Selection

Gupta et al. [48] finds naive clustering of high-dimensional contextual BERT embeddings produces deficient results. An et al. [47] reinforces this theory by surveying embedding model’s clustering ability using Spatial Histograms, which found high-dimensional dynamic SBERT to be least able to cluster when compared to low-dimensional static GloVe models. We argue that by reducing embedding dimensionality and therefore clustering complexity, an increase in clustering performance can be observed.

Established techniques such as Principal Component Analysis (PCA) [49] observe the principal components with maximal variance in an unsupervised methodology. These seek to preserve pairwise distance structures [50] at a local level.

Van der Maaten et al. introduces T-distributed stochastic neighbour embeddings (t-SNE) [51]. The algorithm maps high dimensional elements to a 2 or 3-dimensional representation, whilst preserving distances with neighbouring elements. In contrast to PCA, t-SNE seeks to preserve local distances over global distances [50]. It has extensive use for visualisations of high-dimensional vector spaces. However, t-SNE see detrimental performance when mapping to more than 3 dimensions, as it frequently converges on local minima. This prohibits its use for in clustering description embeddings, because of the limited range of dimensions.

Uniform Manifold Approximation and Projection (UMAP) [50] performs non-linear mappings to arbitrarily lower dimensions, as opposed to t-SNE. The algorithm preserves the global structure, whilst displaying superior time efficiency, enabling scaling to significantly larger datasets, which is vital for the Big Data health care domain. Although, UMAP is stochastic algorithm, it may be initialised with a pre-defined seed to ensure deterministic execution. Superior performance over t-SNE and PCA has been shown when classifying MNIST and Fashion-MNIST datasets [50].

We leverage UMAP’s superior performance and adaptability to map variable embeddings across various dimensions: 10, 50,100, 200 and 300.

#### Clustering Algorithms Selection

K-means clustering is prominent method of vector quantization, introduced by MacQueen et al [52]. Datapoints are assigned to a fixed number of clusters, by minimising intra-cluster distances between centroid and all assigned all datapoints. This process is repeated over a specified number of iterations. The number of iterations can be determined by the Lloyd Expectation Maximisation algorithm [53] or set to a maximum number.

In an unsupervised setting, when the number of domains is not predefined, it is challenging to find the optimal number of clusters. This often necessitates reliance on labour-intensive methods such as visualization and human judgment to infer groupings of variables [54]. Moreover, this approach lacks adaptability in modifying the cluster number; it requires the clusters’ number to be specified beforehand and necessitates a complete re-computation of the model for minor adjustments in hyper-parameters. Hierarchical clustering alleviates this inefficiency.

Hierarchal Agglomerative Clustering (HAC) [55] groups high dimensional embeddings into a hierarchical structure based on any distance information. These can then be truncated at a desired level into distinct clusters. The algorithm is highly flexible with satisfactory performance across any distance metrics, as opposed to centroid and median based algorithms. The algorithm offers significant adaptability over simpler methods such as K-means, by allowing fine granularity adjustments by altering the linkage threshold. Stepwise dendrograms enable the visualisation of hierarchal tree structures for comprehensive analysis of variable similarity irrespective of the linkage threshold. Computationally efficiency is greatly increased for lower linkage threshold by only requiring shallow inspections of the hierarchical tree structure, offering major time reductions when compared to K-means. However, its full space partitioning assumption means that all points must be assigned to a cluster forcing outliers to be assigned to a cluster which affects clusters cohesiveness and decreases harmonisation performance. This inefficiency can be addressed by allowing for some points to be treated as noise and not assigned to clusters.

HDBSCAN [56] extends density-based spatial clustering of applications with noise (DBSCAN) [57] by using clustering hierarchy in addition to allowing for noise points, i.e., outliers, which are not assigned to clusters. Empirical testing demonstrates substantial performance gains over competing algorithms such as OPTICS [58] in the majority of cases. Although due to the algorithms complexity a major computation expenditure is necessary, when compared to K-means, it is still significantly faster than HAC for large datasets.

Density based algorithms, such as DBSCAN, can efficiently identify anomalies in low density region as and discarded them in accordance with a single linkage: minimum number of samples, which dictates the minimum number of neighbouring components to a core point for it be established.

HDBSCAN generalises this with an additional hierarchal minimum cluster size parameter, which states clusters with fewer components are not established and deemed spurious. By forgoing clustering completeness, stronger harmonisations may be achieved. An extension of Prim’s algorithm is used to construct a Minimum Spanning Tree, given density-based groupings, in order to extract the HDBSCAN hierarchy. An optimisation method is used to extract a globally optimal solution from the hierarchal structure [47–51].

#### Clustering Goodness Metrics Selection

Evaluating the goodness of clustering results across various clustering algorithms, hyper-parameters and dimensional mappings has long been seen as a vital issue essential to the success of clustering applications [59]. Clustering validation evaluates the goodness of clustering results [60] without the need for external validation measures such as labelled validation datasets.

Lie et al. [61] reviews 11 metrics and analyses properties such as monotonicity, noise, density, subclusters criteria, in addition to the criteria of compactness and separation. Empirical evidence suggests silhouette score [62] correctly identifies optimal clustering in most cases, however it promotes the merging of nearby subclusters into one for datasets with prominent subclusters, in order to maximise inter-cluster separation. In contrast, S_Dbw [63] satisfies all five aspects, at the expense of computational complexity. However, this property may not be desirable for use with sparse embeddings from SBERT models, as it may prioritise smaller subclusters, dividing semantically similar variables into separate clusters. Nisha et al [64] also promotes the use of silhouette score for evaluating the goodness of clustering. The silhouette score is valued in clustering analysis for its ability to measure both the cohesion within clusters and the separation between them, providing a combined metric that ranges from -1 to 1. It is applicable to various clustering methods without requiring ground truth labels, making it suitable for unsupervised learning scenarios. However, it can be computationally intensive.

We incorporate the Silhouette score goodness of clustering metric due to its favourable qualities [61] and reported performance. The metric computes the pairwise difference between intra-cluster (within cluster) and inter-cluster (between clusters) distances [62].

### Validation Approach

To analyse and validate the performance of the Semantic Search and Semantic Clustering pipelines we create a testing dataset by manually partitioning a set of variables, an appropriate approach when ground truth data absent [47]. We developed validation domains building on the Simpson et al. [5] Delphi Study which identifies 31 domains related to determinants of improved care in multimorbidity. We identified a subset of 12 validation domains relevant to ELSA variable descriptions including: finance; housing; engagement in meaningful activities and social participation; Access to social care, community-based services and other provision; Use of technologies to support individuals at home; Recognition of and support with lifestyle factors; Prescribing and medication management; Enhanced support from family and other informal carers; Person-centred and holistic care; Supporting self -management of conditions; Support with daily living and independent living; and Environmental factors and wider social determinants of health. A random sample of 2000 variables from the ELSA dataset were taken, and manually labelled with 12 validation domains to create a test set for comparison. Manual comparison is performed only using the description of variable, and no other external information, allowing for comparison between human and automated pipelines performance.

For the Semantic Search pipeline evaluation, the resulting cosine similarity score for each variable are evaluated using the AUC metric [65], calculating the area under the receiver operating characteristic (ROC) curve. This ensures performance is measured for a given validation domain and search phrase, irrespective of the chosen similarity threshold, by comparing against the labelled test set.

For Semantic Clustering pipeline evaluation, we firstly use Silhouette score [62]to converge on optimal set of clusters and then use the V-measure[66,67] to evaluate clustering performance against the test set. Standard pairwise comparison is not possible as the arbitrary number of clusters is not equal to the fixed number of 12 validation domains in our test set, requiring an alternative approach. Therefore, for a given domain the cluster with maximum V-measure is assumed to match that domain. To quantify harmonisation performance across multiple embedding dimensions and clustering algorithms, a mean of the maximal v-measures is taken across all domains to enable thorough comparison. Boltužić et al [67] utilised the V-measure metric [66], measuring a harmonic mean of homogeneity and completeness, which are more desirable aspects of clustering than accuracy. As opposed to precision and recall, V-measure is not influenced by incomplete clustering, where some elements are not clustered. The measure is independent of the dataset or clustering algorithm utilised and it vital to note that it favours more coherent incorrect samples. Similar measures such Q2 [70], are dependent of the number of clusters and do not explicitly calculate completeness. V-measure [71] is invariant to the number of clusters. Empirical evidence demonstrates effective evaluation of highly dimensional TF-IDF vectors [66], as well as Transcriptomic Data for Breast and Lung Cancer [72] using V-measure.

## Results

### Semantic Search Evaluation

Table 1 captures the accuracy of variable selection using AUC for Semantic Search on the test set of 12 domains, each described by a search phrase, including 2000 variables.

**Table 1:** Area under the curve metrics across all sentence embedding models tested, when matching a user generated search phrase to manually labelled validation domains.

| Domain<br>(Simpson et al. Table 5 care need determinant number) | Search Phrase | AUC |  |  |  |
| --- | --- | --- | --- | --- | --- |
|  |  | MiniLM | MPNet | BioLink BERT | T5-XXL |
| Finance/ financial assistance (18) | Finance, inherit, insurance or benefits | 0.828 | 0.800 | 0.601 | **0.873** |
| Housing/accommodation that meets individual's needs (25) | House, mortgage, or property | **0.920** | 0.883 | 0.673 | 0.851 |
| Able to engage in meaningful activities and social participation (22) | Current job or Retirement | 0.823 | **0.889** | 0.659 | 0.838 |
| Access to social care, community-based services and other provision (7) | Formal help received such as Nurse or Doctor | 0.910 | 0.853 | 0.697 | **0.911** |
| Use of technologies to support individuals at home (31) | Technology devices, Aids, or cars | **0.931** | 0.891 | 0.633 | 0.822 |
| Recognition of and support with lifestyle factors (23) | Diet, Exercise, Alcohol and Smoking | 0.793 | **0.850** | 0.685 | 0.828 |
| Prescribing and medication management (14) | Medication, Drugs taken and tablets | **0.993** | 0.988 | 0.606 | 0.987 |
| Enhanced support from family other informal carers (27) | Informal help received | 0.873 | 0.886 | 0.703 | **0.906** |
| Person-centred and holistic care (1) | Measurements of Blood and other bodily functions | **0.930** | 0.928 | 0.699 | 0.879 |
| Supporting self - management of conditions (12) | How did you feel or emotions | **0.946** | 0.911 | 0.698 | 0.851 |
| Support with daily living and independent living (16) | Received help with daily tasks | 0.933 | 0.926 | 0.698 | **0.946** |
| Environmental factors and wider social determinants of health (21) | Environment outdoors | 0.914 | 0.937 | 0.698 | **0.970** |
|  | **Mean (SD):** | **0.899 (0.056)** | 0.895 (0.046) | 0.671 (0.036) | 0.888 (0.053) |

We observe that the performance of the domain-specific embedding BioLinkBERT is inferior in comparison to other generalised embeddings. The remaining general embedding models MiniLM, MPNet and T5-XXL have comparative performance, however, MiniLM exhibited the highest AUC score (M=0.899 SD=0.056), as well as having smallest model size. Smaller models require less memory and computational power and generally load and execute faster. Therefore, MiniLM can be assumed the best performant model. Interestingly, SBERT exhibited Named Entity Recognition (NER) abilities, linking entities within similar semantic use cases. Tobacco products such as “Paan Masala” and “Bidi” where harmonised within the same lifestyle domain.

### Semantic Clustering Evaluation Results

Table 2 captures the variables grouping accuracy of Semantic Clustering on the test set using the three clustering algorithms under assessment.

**Table 2:** The highest Maximum V-measure averaged over each validation domain across each clustering algorithm. UMAP projections of MiniLM variable description embeddings were clustered using: K-means (centroids = 77), Hierarchal Agglomerative Clustering (linkage = 0.01), HDBSCAN (minimum samples = 20 and minimum cluster size = 20).

| Clustering Algorithm | Dimensions | Allocated Clusters | Silhouette | Mean Max V-measure (SD) |
| --- | --- | --- | --- | --- |
| K-means (77) | 50 | 76 | 0.662 | 0.223 (0.125) |
| HAC (0.01) | 50 | 3 | 0.685 | 0.079 (0.110) |
| HDBSCAN<br>(20,20) | 300 | 25 | **0.817** | **0.237 (0.157)** |

**Table 3:** Time taken in seconds for the encoding of 2000 variable descriptions using MiniLM and clustered using: K-means (centroids = 77), Hierarchal Agglomerative Clustering (linkage = 0.01), HDBSCAN (minimum samples = 20 and minimum cluster size = 20).

| Clustering Algorithm | Encoding Time (s) | Clustering Time (s) | Evaluation Time (s) | Total Time (s) |
| --- | --- | --- | --- | --- |
| K-means (77) | 3.720 | 5.286 | 3.675 | 12.681 |
| HAC (0.01) | 3.720 | 0.117 | 0.160 | 3.997 |
| HDBSCAN (20,20) | 3.720 | 1.529 | 0.824 | 6.073 |

For the evaluation of Semantic Clustering, we adopt the best Semantic Search embedding model MiniLM due to its optimal performance and low computational requirements. The original 384-dimensional embedding is reduced using UMAP into a range of dimensions: 10, 50, 100, 200, 300. Silhouette score was used to select optimal clustering, enabling thorough hyper-parameter tuning of the algorithms. Table 2 displays the Mean Max V-Measure (MMV) of the optimal clustering using K-means, HAC or HDBSCAN. We find HDBSCAN to produce superior results allocating 25 clusters with a maximum Silhouette score of 0.817 and maximum MMV of 0.237 (SD=0.157), when a 20 minimum cluster size of and 20 minimum number of samples is used. Performance is in line with comparable study by Boltužić et al [67]. A minor reduction to 300 dimensional embeddings was optimal for this task, indicating HDBSCAN’s superior performance in high dimensional spaces when compared to HAC and K-means. Both K-means and HAC gave optimal clustering with 50 dimensional embeddings, indicating difficulty in clustering high-dimensional vector spaces.

HAC was unable to discriminate clusters when applied after UMAP dimensionality reduction. When using the lowest linkage value of 0.01, only a single homogenous cluster was allocated for 200 and 300 dimensional embeddings. When analysing HAC dendrograms, we observed that neighbouring clusters are semantically dis-similar [64].

## Discussion

We observe high accuracy of the Semantic Search pipeline, with mean AUC across the 12 domains of 0.899 (SD=0.056) for the best performing embedding model MiniLM. The Semantic Clustering pipeline performance is on par with leading implementations in argumentation mining [67], with a mean maximum V-measure of 0.237 (SD=0.157).

Considerable time and resource savings are accomplished by employing the automated pipelines, both for Semantic Search and Semantic Clustering. The execution times of the longest running pipeline, Semantic Clustering, are shown in **Error! Reference source not found.**, with fastest configuration being with the HDBSCAN clustering method, only taking 4.85 seconds to encode and cluster 2000 variable descriptions. Restricting the pipeline to MiniLM and HDBSCAN algorithms, tuning was performed using a Grid Search across 5 UMAP dimensions and 13 different HDBSCAN minimum cluster sizes. 65 iterations were processed within 510 seconds.

Similarly, Semantic Search across all ELSA variables is also performed in seconds. In contrast, manual labelling of 2000 variables took approximately 16 hours, costing significant human resources.

Extrapolating this to 22,402 unique variables, a manual labelling of entire dataset would take 176 person-hours. In our experiment, the speed of automated variables clustering and assigning to clusters (approximately 245 variables per minute) is over 100 times faster than the speed of manual variables labelling (approximately 2.1 variables per minute). Using ML technologies can dramatically aid data harmonisation for Big Data datasets, catalysing future health data science research.

Currently it is not possible to directly compare our validation domains to benchmark datasets as they do not exist for the study and curation of MLTCs and social care needs. However, we can assess the differences in the approaches for applying techniques for datasets incorporating other domains.

Sui X et al. opt to train classification models [73], necessitating the use of ground truth training sets for the target domains. Our approach avoids this requirement by using unsupervised methods.

Landthaler et al. [74] extends text search for legal documents using a static Word2Vec embeddings in conjunction with a t-SNE visualisation. Successfulempirical grouping of sentences it shown, but no performance evaluation is provided. Boltužić et al [67] performs a small-scale STS task upon textual online debate forums, identifying prominent arguments in an unsupervised manner, with HAC and simpler Skip-gram methods. This achieves V-measure results in the range of 0.15 to 0.30 with an average of 0.233, which is in line with our more sophisticated SBERT embeddings methodology with an MMV of 0.237 (SD=0.157) when using HDBSCAN. The study uses a dataset of 3014 sentences, similar to our use of 2000 randomly selected ELSA variables (from the over 120,000 ELSA variables).

Boltužić et al [67] performs a small-scale STS task upon textual online debate forums, identifying prominent arguments in an unsupervised manner, with HAC and simpler skip-gram methods. This achieves V-measure results in the range of 0.15 to 0.30 with an average of 0.233, which is in line with our more sophisticated SBERT embeddings methodology with an MMV of 0.237 (SD=0.157) when using HDBSCAN. The study uses a dataset of 3014 sentences, similar to our use of 2000 randomly selected ELSA variables (from the approximately 120,000 ELSA variables).

It is important to note that any such model performance is subjective and conditional upon the phrase inputted. As the ELSA dataset features minimal specialised medical terminology, generalised models such as MiniLM trained on a general English language corpus exhibit increased performance. However other specialised datasets with domain specific terminology in variable descriptions, could see substantial improvements with domains specific models, such as BioLinkBERT.

Semantics-aware search and clustering discussed in this manuscript are general and applicable to other electronic healthcare data. However, to increase the usability of Semantic Clustering further efforts need to be made to increase human interpretability of the output clusters. Visualisation tools such as ClusterVision [54] could assist with the interpretation of high-dimensional embedding clusters, enabling identification of embedding semantic misidentifications and biases.

## Conclusions

In recent years observational retrospective clinical studies have emerged as valuable alternatives to traditional clinical trials offering cost-effectiveness and efficiency while still generating valid results. The availability of cohort and routine databases, such as ELSA, CPRD, and SAIL, has been a significant catalyst of this trend by providing access to vast amounts of data, known as big data in the field of data science. Leveraging this big data, however, requires substantial efforts in harmonising individual source datasets and curating study data, as the current process relies on manual and labour-intensive methods.

In this manuscript we discussed the research and validation of AI technologies, particularly in the areas of natural language processing (NLP) and unsupervised ML, to streamline the harmonisation and curation of datasets for observational studies using healthcare big data sources. We explored the latest advancements in NLP and unsupervised ML techniques needed for the development of automated tools for the harmonisation process.

We proposed two pipelines: Semantic Search for domain-relevant variable identification and Semantic Clustering for identifying semantically similar variables. These pipelines combine state -of-the-art AI algorithms, such as MiniLM pretrained Sentence-BERT model for semantics aware text embedding, UMAP for dimensionality reduction and HDBSCAN for clustering. The performance of these pipelines was evaluated using the ELSA database .

Our results demonstrate high accuracy in Semantic Search, achieving an AUC of 0.899, while Semantic Clustering exhibited performance comparable to leading implementations in other domains, with a V-measure of 0.237 (SD=0.157). Importantly, our automated tools significantly reduced the time and resources required for data harmonisation and curation compared to manual approaches.

Our study findings underscore the potential of AI technologies, such as NLP and unsupervised ML, in automating the harmonisation and curation of big data for clinical research. By establishing a robust technological foundation, we pave the way for the development of automated tools that streamline the process, enabling researchers to leverage big data more efficiently and effectively in their studies.

## Declarations

### Data availability

The research data was made available to access through ELSA and as such, our study data cannot be made available for access.

## Acknowledgements

We express our gratitude to Hajira Dambha-Miller for her leadership as the principal investigator of project NIHR202637. We also extend our thanks to all the project team members. Without their participation this work would not have been possible.

## Abbreviations

AI: Artificial Intelligence
BERT: Bidirectional Encoder Representations from Transformers
DBSCAN: density-based spatial clustering of applications with noise
EHR: Electronic Healthcare Records
ELSA: English Longitudinal Study of Ageing
HSE: Health Survey for England
HAC: Hierarchal Agglomerative Clustering
HDBSCAN: Hierarchal density-based spatial clustering of applications with noise
ICU: Intensive Care Units
ML: Machine Learning
MAP: Mean Average Precision
MMV: Mean Max V-Measure
MLTC: Multiple Long-Term Conditions
NLP: Natural Language Processing
PCA: Principal Component Analysis
STS: Semantic Textual Similarity
SBERT: Sentence-BERT
SemDHP: Semantic Data Harmonisation Pipeline
SNLI: Stanford Natural Language Inference
t-SNE: T-distributed stochastic neighbour embeddings
T5: Text-to-Test Transfer Transformer
UMLS: Unified Medical Language System
UMAP: Uniform Manifold Approximation and Projection
USE: Universal Sentence Encoders

